# Supporting the implementation of Osteoarthritis Management Programs in low-and middle-income settings: Understanding clinician training outcomes alongside perceived program barriers and facilitators in Malawi

**DOI:** 10.1101/2025.02.07.25321791

**Authors:** Nonhlanhla S. Mkumbuzi, Esther Jiya, Enock M. Chisati, Joanne L. Kemp, Christian Barton, Allison Ezzat

## Abstract

**Objective:** To examine Malawian physiotherapists’ knowledge and beliefs about Osteoarthritis (OA) and their perceived capabilities to deliver an Osteoarthritis Management Program (OMP) to people with knee and hip OA; and to identify OMP barriers and facilitators in Malawi.

**Design:** Two-phased mixed methods formative evaluation

**Methods:** Phase 1: Malawian physiotherapists participated in the GLA:D^®^ Australia training course and answered quantitative pre-and-post course questions that were descriptively summarized, and analysed using McNemar’s test, where appropriate. Phase 2: semi-structured focus groups generated qualitative data that were thematically analysed and mapped to the Consolidated Framework for Implementation Research. Mixed methods data were integrated through triangulation.

**Results:** Eleven Malawian physiotherapists [9 (82%) female, 10 (91%) with 5–10 years clinical experience] participated. Post training course, participants’ knowledge of OA management increased regarding the benefits of therapeutic exercise (p=0.002), importance of weight management (p=0.004), and acceptable symptoms profile (p=0.008). Participants’ confidence and beliefs in managing knee and hip OA also increased. Implementation barriers included program costs, current medical management of OA with painkillers, and infrastructure challenges. Implementation facilitators included the content and organisation of GLA:D^®^, the ability to adapt the program, and OA awareness and education among other health professionals.

**Conclusion:** Knowledge, confidence, and beliefs in managing knee and hip OA improved post GLA:D^®^ training in Malawian physiotherapists. Increasing education of physiotherapists, other health professionals and the general public about evidence-based OA management and making contextual adaptions to the GLA:D^®^ training and program structure may facilitate future implementation of OMP, such as GLA:D^®^, in low-and-middle income countries.

## INTRODUCTION

Non-communicable diseases account for 61% of global disability adjusted life years, with osteoarthritis (OA) affecting 595 million people globally, thus contributing substantially to this burden.^1,2^ Knee and hip OA cause debilitating joint pain and stiffness, activity limitations, and loss of productivity^3–5^ at the individual level, alongside significant economic burden at community and healthcare systems levels worldwide.^6^ All high quality clinical guidelines for the management of hip and knee OA recommend patient education and exercise therapy as critical first line interventions.^7,8^ Exercise-therapy has been found to reduce pain and improve function and quality of life, while also being cost-effective and may allow some patients to delay or avoid joint replacement surgery. ^9–12^ Globally, many individuals with OA, including those who live in low and middle income countries (LMICs), do not have access to exercise therapy or patient education.^6^

The diagnoses of hip and knee OA are increasing in LMICs^1,13^. Between 2010 and 2017, the prevalence of OA in South Africa, Brazil, and Nepal rose by 9, 14, and 20%, respectively.^2^ Recent studies in Nigeria, Democratic Republic of Congo, Egypt, and Cameroon also reported OA prevalence of up to 37%.^13,14^ However, the majority of first line interventions developed and evaluated for the management of OA have been conducted in high-income countries.^15–18^ Hence, little is known about how to adapt and implement evidence-based first line interventions developed in high or upper middle income settings into low income settings for the management of knee and hip OA.

The Good Life with osteoArthritis from Denmark (GLA:D^®^) program is an evidence-based hip and knee Osteoarthritis Management Program (OMP) that consists of supervised group education and exercise-therapy program.^15,16^ The program trains physiotherapists to deliver high quality education and individualized exercise-therapy to patients across the full spectrum of OA severity and has been implemented in more than ten high income countries worldwide.^16^ Program evaluation of GLA:D^®^ in Australia found that 75% of people report clinically meaningful improvement in pain or quality of life, and 3 of 4 people wanting surgery prior to participation did not have surgery and no longer desired surgery at 12-months follow up.^19,20^ Despite these demonstrated clinical benefits of the GLA:D^®^ program in high income countries, it has not been implemented in any LMICs.^15^ It is unknown how it could be contextually adapted for implementation in a low income country and if GLA:D^®^ would be clinically effective or considered culturally appropriate in these settings.^21–23^

The aim of the present study was two-fold. Firstly, to examine Malawian physiotherapists’ knowledge and beliefs about OA and their perceived capabilities to provide an OMP to people with knee and hip OA. Secondly, to identify barriers and facilitators of implementing the physiotherapist training and the OMP for people with OA in Malawi. This work will generate new knowledge that could contribute to the future implementation of OMP in Malawi and other LMICs.

## METHODS

### Study design

A two-phased mixed methods formative evaluation was conducted to develop a comprehensive understanding of the feasibility of future implementation of the GLA:D^®^ program in Malawi from the perspective of physiotherapists. A mixed methods design is highly suited to the examination of complex phenomena and hard-to-measure constructs as it allows a research question to be approached from multiple perspectives.^24^ This convergent parallel design collected quantitative (survey) and qualitative (focus group) data simultaneously before comparing and relating the findings together.

In phase 1, Malawian physiotherapists participated in the GLA:D^®^ Australia training course and answered quantitative pre-and-post course questions to understand their knowledge and beliefs about OA management. In phase 2, physiotherapists perspectives on factors influences the implementation of GLA:D^®^ in Malawi was explored using semi-structured focus groups. These two phases produced complementary results which were triangulated to produce a deeper, more complete understanding of the challenges and opportunities associated with implementation of GLA:D^®^ in Malawi.

Study reporting was guided by the COnsolidated criteria for REporting Qualitative research (COREQ) checklist^25^ and Good reporting of a mixed methods (GRAMM) study checklist^26^ (Supplementary Tables 4 and 5). Ethical approval was obtained from the College of Medicine Research and Ethics Committee, Kamuzu University of Health Sciences, Malawi (P.08/22/3707) and La Trobe University Human Research Ethics Committee, Australia (REF: HEC23003).

### Participants

Malawian physiotherapists were purposively recruited by a local research team member (EC) via the Malawian Physiotherapy Association networks, Kamuzu University of Health Sciences, social media, and word of mouth. Sampling sought to engage a diverse group of physiotherapists from a variety of practice settings (public and private clinic or hospital) with a range of clinical experience and professional development training. Eligibility criteria were that physiotherapists had to be registered to practice in Malawi, have a minimum 50% of their workload dedicated to patient care, and regularly treat patients with hip or knee OA. Physiotherapists with previous GLA:D^®^ training were ineligible. A pragmatic sample size of minimum ten participants were targeted to meet the aims of this formative evaluation.

### GLA:D® Training course

Participants engaged in an online, modified version of the GLA:D^®^ Australia clinician training course delivered by members of the GLA:D^®^ Australia leadership team (JK, CB, AE). Participants attended together at the Rehabilitation Sciences Department of Kamuzu University of Health Sciences in Blantyre, Malawi. The course was delivered in 4 hour sessions over three consecutive days. Briefly, the course included lectures and discussions on OA pathophysiology, diagnosis, and management, with an emphasis on first line care delivered by physiotherapists (education, exercise). Additionally, participants engaged in educational role playing scenarios and practiced the standardized GLA:D^®^ exercises, including how to modify exercises based on patient’s individual capacity and equipment access.^19^

### Phase 1: Quantitative Data Collection & Analyses

Demographic and practice details were collected from participants. Immediately before and after the GLA:D^®^ training course, participants answered questions evaluating their OA knowledge and management (21 questions); self-efficacy and beliefs in their ability to provide first line care for hip and knee OA (9 questions); beliefs regarding evidence for different interventions for hip and knee OA (15 questions); and confidence in abilities to deliver different interventions for hip and knee OA (14 questions). After the training course, participants answered an additional 16 questions on GLA:D^®^ policies and procedures and 6 GLA:D^®^ training course evaluation questions.

Demographic and practice characteristics were summarized descriptively using frequency (percentage). Responses to all further questions were also summarized descriptively using frequency (percentage) or median (Q1;Q3), as appropriate, and percent change between pre and post GLA:D^®^ course was reported. McNemar’s test for paired, non-parametric data examined significant differences between pre-and-post GLA:D^®^ training course questions (p < 0.05). This included the proportion of correct responses on OA knowledge and management (overall and for each question), self-efficacy in ability to provide first line care (dichotomized into strongly agree and agree versus neutral, disagree, and strongly disagree); beliefs about evidence for interventions (dichotomized into strongly supports and supports versus unclear and doesn’t support), and confidence in delivering different interventions (dichotomized into very confident and confident versus average, below average, and not confident at all).

### Phase 2: Qualitative Data Collection & Analysis

This phase was underpinned by an interpretive descriptive paradigm^27^ and informed by the Consolidated Framework for Implementation Research (CFIR).^28^ CFIR is a widely used determinants framework in global implementation research to guide the exploration of factors that influence program implementation. It has been used and evaluated as compatible for application in LMICs largely due to its comprehensiveness and flexibility.^29^ In this study, we used the recently updated version of CFIR, which includes 48 constructs within five domains.^28^

Immediately following the GLA:D^®^ training course, participants engaged in a 2 hour semi-structured focus group via videoconference to explore factors (barriers and facilitators) influencing the implementation of GLA:D^®^ in Malawi. The focus group involved three small group break out discussions interspersed with large group shared summary discussions. The topic guide was developed by the research team and consisted of open ended questions organized in the CFIR domains (Appendix A – Focus Group structure and questions). The topic guide was pilot tested with two Malawian physiotherapists not participating in the study and no changes were made. Experienced qualitative researchers facilitated the focus group [AE (women) and CB (man) both physiotherapists with PhDs, clinical experience with the GLA:D^®^ program, and extensive qualitative training (>10 years)]. A third researcher also contributed to the analysis process [EJ (woman)]. None of the researchers had any prior relationship with the participants. Participants were aware that the research team included a mix of GLA:D^®^ Australia leadership members and local African researchers with an interest in improving management of knee and hip OA in LMICs.

Focus groups were recorded and transcribed verbatim. Data were managed in NVivo qualitative data analysis software (QSR International Pty Ltd). Qualitative analysis used a multi-step inductive thematic approach.^27^ Throughout analysis, trustworthiness was enhanced by detailed reflective memoing after the focus group; prolonged engagement with data; and multiple, in-depth discussions of emerging themes by three authors.

Initially, two researchers (AE, EJ) independently reviewed and inductively coded transcripts line-by-line. They met multiple times to iteratively discuss, refine, and come to a shared understanding of a coding framework. Subsequently, they consolidated codes into subthemes and deductively mapped these to the five broad CFIR domains (Innovation, Outer Setting, Inner Setting, Individuals, Implementation Process). Authors AE and CB met three times to reflect on and discuss the subthemes in relation to the CFIR framework. Next, one researcher (AE) further mapped the sub-themes to the 48 individual CFIR constructs within the five domains and interpreted each as an implementation barrier or facilitator. Barriers were conceptualized as themes where focus group participants perceived a negative impact on GLA:D^®^ program implementation, whereas facilitators were conceptualized as having a positive impact on GLA:D^®^ program implementation. The framework of CFIR constructs used for this final stage of analysis was shared with researcher EJ who independently repeated the mapping and classification of themes. All three researchers participated in final discussions to confirm data alignment with CFIR constructs, reach consensus on the classification of each sub-theme as a barrier or facilitator, and articulate a final theme for each CFIR domain.

### Data Integration

After quantitative and qualitative data were analysed independently, they were triangulated through team discussion for a richer, more in-depth understanding of implementation of GLA:D in Malawi.

## RESULTS

Eleven Malawian physiotherapists from a variety of practice settings participated in the study (**Table 1**).

**Table 1:** Demographic and practice characteristics of Malawian physiotherapists (n=11)

| Characteristic | n (%) |
| --- | --- |
| **Gender** |  |
| Man | 2 (18) |
| Woman | 9 (82) |
| **Location of practice\*** |  |
| Remote | 2 (18) |
| Rural | 8 (73) |
| Urban | 7 (64) |
| **Practice setting\*** |  |
| Private clinic or hospital | 7 (64) |
| Public in-patients or out-patients | 2 (18) |
| Community | 5 (46) |
| Aged care | 7 (64) |
| Sporting team | 4 (36) |
| **Length of practice** |  |
| 5 –10 years | 10 (91) |
| 10 –15 years | 1 (9) |
| **Area of practice** |  |
| Musculoskeletal | 1 (9) |
| A mixture of musculoskeletal and others | 10 (91) |
| **Have you ever completed any post-graduate training?** |  |
| Yes | 2 (18) |
| No | 9 (82) |
| **Aware of any OA clinical practice guidelines** |  |
| No | 11 (100) |

\*Participants could select more than one response

### Phase 1: Quantitative results

Participants’ OA knowledge and management pre-and-post training course is summarized in **Table 2**. Briefly, participants’ knowledge of OA management increased regarding the benefits of therapeutic exercise (p=0.002), importance of weight management (p=0.004), and acceptable symptoms profile (p=0.008).

**Table 2:**
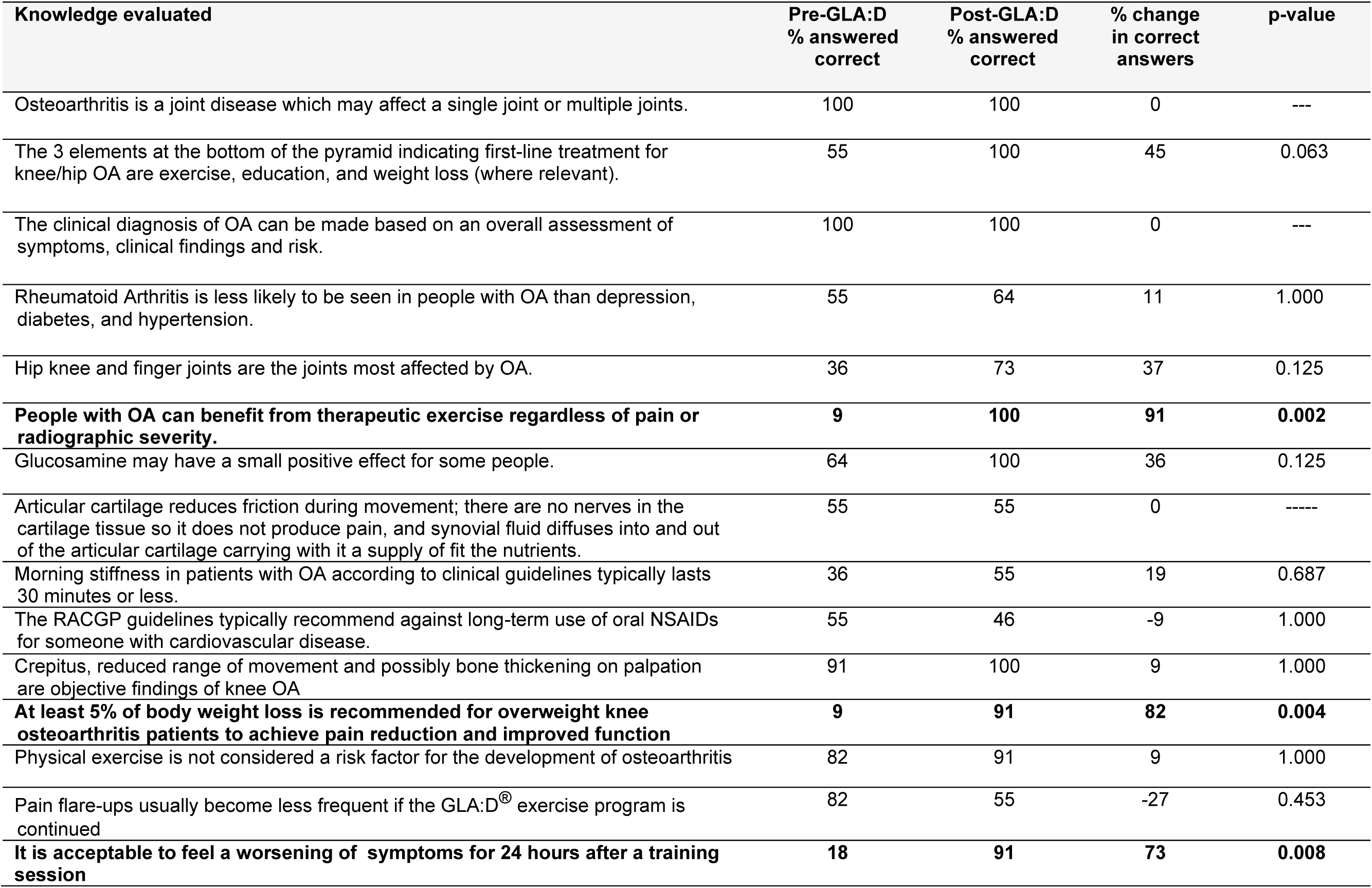
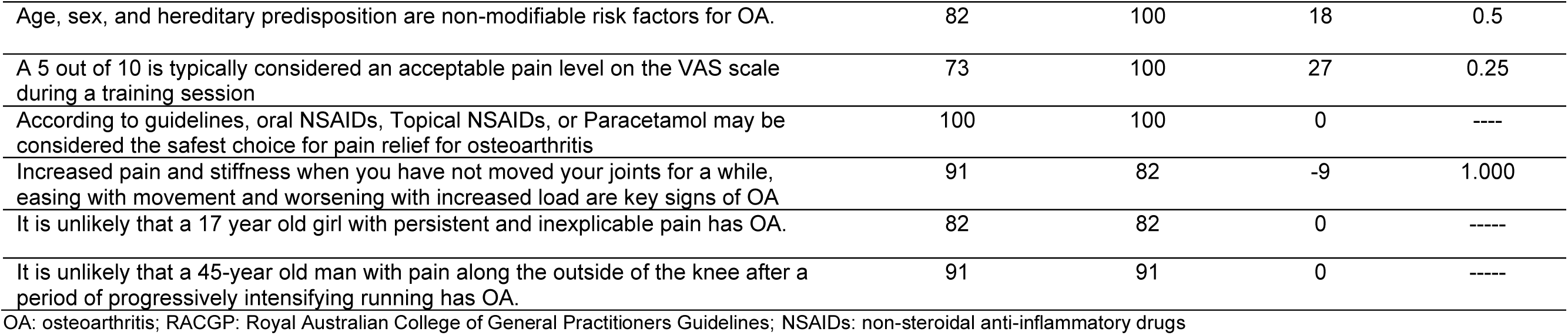
Pre-and post-GLA:D^®^ training course OA knowledge and management among Malawian physiotherapists (n=11)

| Knowledge evaluated | Pre-GLA:D<br>% answered<br>correct | Post-GLA:D<br>% answered<br>correct | % change<br>in correct<br>answers | p-value |
| --- | --- | --- | --- | --- |
| Osteoarthritis is a joint disease which may affect a single joint or multiple joints. | 100 | 100 | 0 | --- |
| The 3 elements at the bottom of the pyramid indicating first-line treatment for knee/hip OA are exercise, education, and weight loss (where relevant). | 55 | 100 | 45 | 0.063 |
| The clinical diagnosis of OA can be made based on an overall assessment of symptoms, clinical findings and risk. | 100 | 100 | 0 | --- |
| Rheumatoid Arthritis is less likely to be seen in people with OA than depression, diabetes, and hypertension. | 55 | 64 | 11 | 1.000 |
| Hip knee and finger joints are the joints most affected by OA. | 36 | 73 | 37 | 0.125 |
| **People with OA can benefit from therapeutic exercise regardless of pain or radiographic severity.** | **9** | **100** | **91** | **0.002** |
| Glucosamine may have a small positive effect for some people. | 64 | 100 | 36 | 0.125 |
| Articular cartilage reduces friction during movement; there are no nerves in the cartilage tissue so it does not produce pain, and synovial fluid diffuses into and out of the articular cartilage carrying with it a supply of fit the nutrients. | 55 | 55 | 0 | ----- |
| Morning stiffness in patients with OA according to clinical guidelines typically lasts 30 minutes or less. | 36 | 55 | 19 | 0.687 |
| The RACGP guidelines typically recommend against long-term use of oral NSAIDs for someone with cardiovascular disease. | 55 | 46 | -9 | 1.000 |
| Crepitus, reduced range of movement and possibly bone thickening on palpation are objective findings of knee OA | 91 | 100 | 9 | 1.000 |
| **At least 5% of body weight loss is recommended for overweight knee osteoarthritis patients to achieve pain reduction and improved function** | **9** | **91** | **82** | **0.004** |
| Physical exercise is not considered a risk factor for the development of osteoarthritis | 82 | 91 | 9 | 1.000 |
| Pain flare-ups usually become less frequent if the GLA:D® exercise program is continued | 82 | 55 | -27 | 0.453 |
| **It is acceptable to feel a worsening of symptoms for 24 hours after a training session** | **18** | **91** | **73** | **0.008** |
| Age, sex, and hereditary predisposition are non-modifiable risk factors for OA. | 82 | 100 | 18 | 0.5 |
| A 5 out of 10 is typically considered an acceptable pain level on the VAS scale during a training session | 73 | 100 | 27 | 0.25 |
| According to guidelines, oral NSAIDs, Topical NSAIDs, or Paracetamol may be considered the safest choice for pain relief for osteoarthritis | 100 | 100 | 0 | ---- |
| Increased pain and stiffness when you have not moved your joints for a while, easing with movement and worsening with increased load are key signs of OA | 91 | 82 | -9 | 1.000 |
| It is unlikely that a 17 year old girl with persistent and inexplicable pain has OA. | 82 | 82 | 0 | ----- |
| It is unlikely that a 45-year old man with pain along the outside of the knee after a period of progressively intensifying running has OA. | 91 | 91 | 0 | ----- |

OA: osteoarthritis; RACGP: Royal Australian College of General Practitioners Guidelines; NSAIDs: non-steroidal anti-inflammatory drugs

Participants’ confidence and beliefs in managing knee and hip OA increased post GLA:D^®^ training are summarised in **Figure 1**. Pre-course, 54% strongly agreed or agreed that they have the skills, 45% strongly agreed or agreed have been trained, and 54% strongly agreed or agreed know how to deliver exercise and education to people with OA based on clinical guidelines. Comparatively, post-course the proportions of physiotherapists who agreed or strongly agreed increased to 100% for each of these three questions, with one being a statistically significant change (p=0.031).

**Figure 1:**
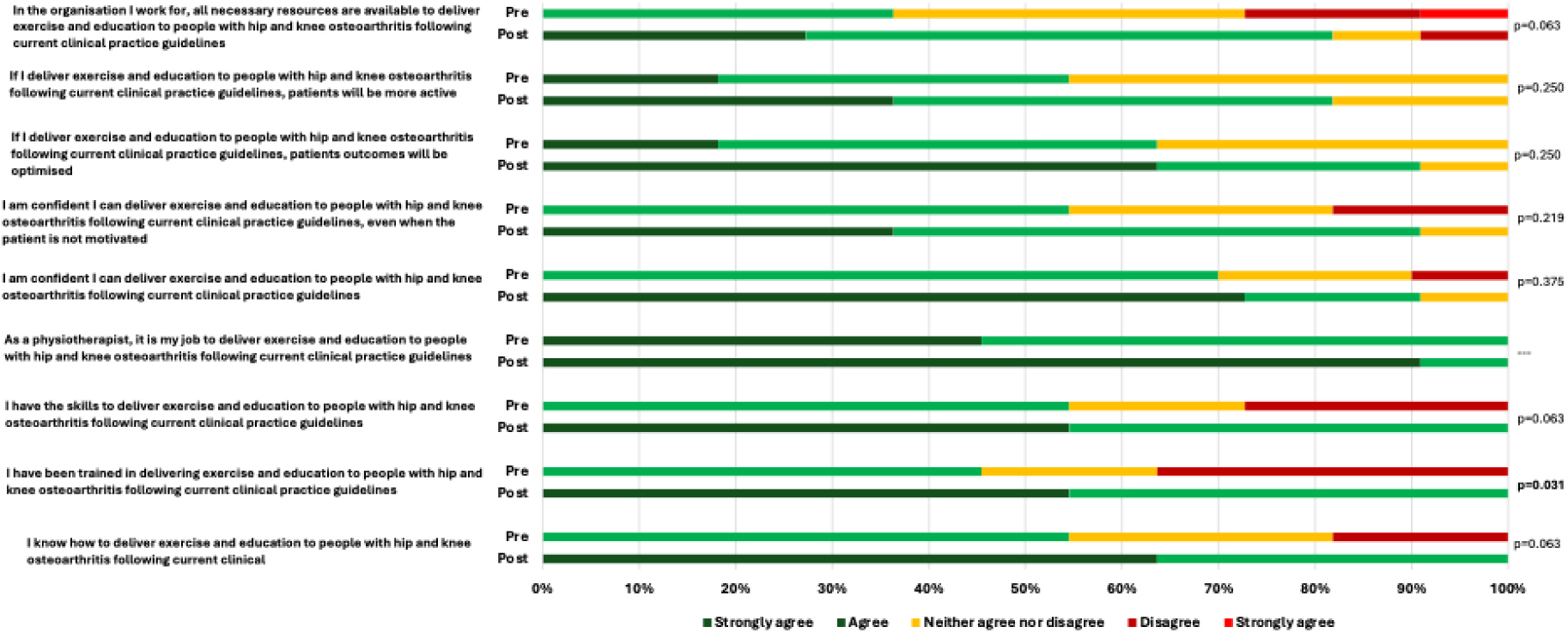
Malawian physiotherapists’ pre-and post-GLA:D^®^ training knowledge and confidence in delivering the GLA:D^®^ program (n=11)

Changes in participants’ beliefs about current evidence for any intervention were not statistically significant, including first line care (exercise therapy, patient education, and weight management) between pre and post training (Table 3; p>0.05).

**Table 3:** Malawian physiotherapists’ pre-and post-GLA:D^®^ training beliefs about current evidence for interventions for hip and knee osteoarthritis (n=11)

|  | Hip OA |  |  | Knee OA |  |  |
| --- | --- | --- | --- | --- | --- | --- |
|  | Evidence strongly supports or supports |  | P value | Evidence strongly supports or supports |  | P value |
|  | Pre-training % | Post-training % |  | Pre-training % | Post-training % |  |
| Exercise therapy | 100 | 100 | - | 100 | 100 | - |
| Patient education | 100 | 100 | - | 100 | 100 | - |
| Weight management | 100 | 100 | - | 100 | 100 | - |
| Electrotherapy | 64 | 36 | 0.250 | 64 | 46 | 0.250 |
| NSAIDs | 82 | 82 | 1.000 | 73 | 82 | 1.000 |
| Opioids | 46 | 64 | 0.625 | 73 | 55 | 0.500 |
| Cortisone injection | 64 | 82 | 0.625 | 64 | 82 | 0.500 |
| Arthroscopy | 46 | 18 | 0.375 | 37 | 46 | 0.625 |
| Joint replacement | 73 | 46 | 0.250 | 82 | 64 | 0.500 |

NSAIDs: non-steroidal anti-inflammatory drugs

Physiotherapists’ confidence in delivering exercise-therapy and general patient education improved, but changes were not statistically significant (Figure 2; p>0.05). Additional questions on specific types of education (physical activity, self-management, pain medication) also shifted towards increased confidence, although less so for questions related to education about benefits and risks of total joint replacement and arthroscopy with 45% and 54% reporting below average confidence, respectively (Figure 2) (additional data in Supplementary Table 2).

**Figure 2:**
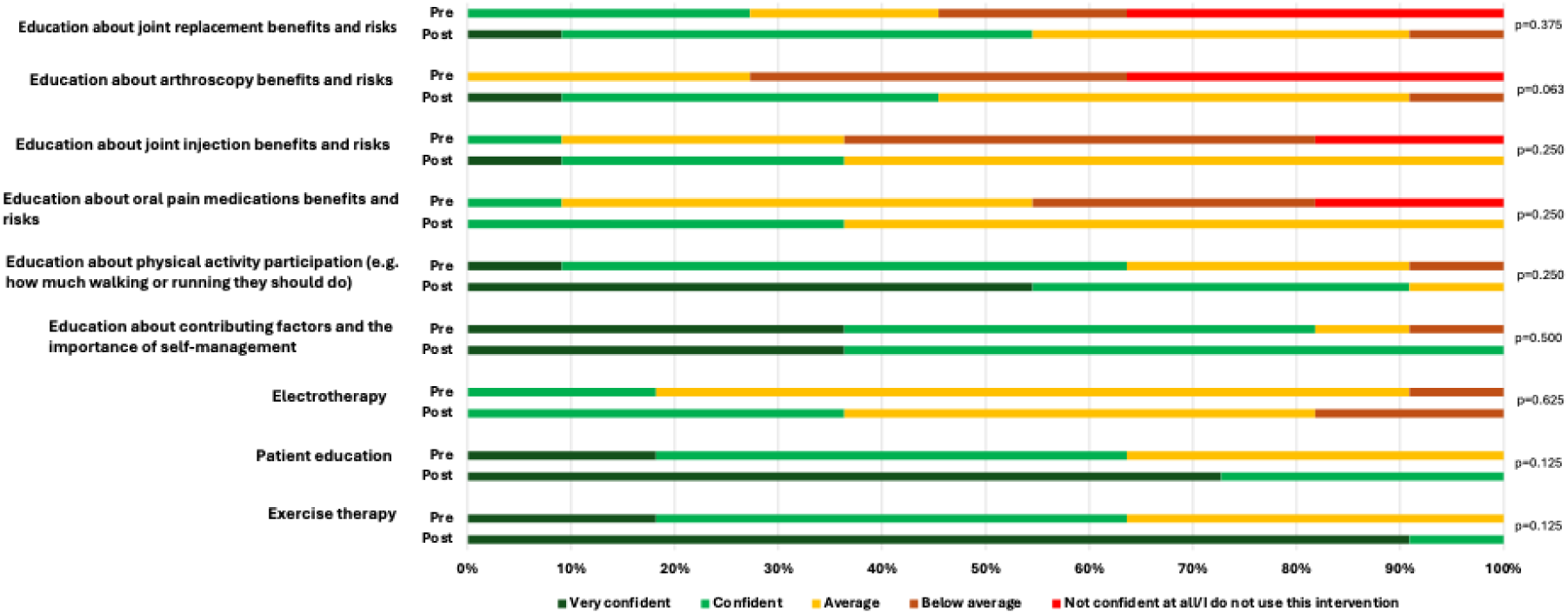
Malawian physiotherapists’ pre-and post-GLA:D^®^ training confidence in delivering interventions for hip and knee osteoarthritis (n=11)

After training, knowledge of GLA:D^®^ procedures and practices was high with a median score of 91% (73, 100) (Supplementary Table 3). All (100%) participants understood the overall objectives of neuromuscular exercises and all participants considered themselves ready to deliver the GLA:D^®^ program to patients.

### Phase 2: Qualitative Results

Barriers and facilitators to GLA:D^®^ implementation in Malawi aligned with all five CFIR domains and were mapped to 18 specific constructs and are summarized in Table 4 alongside illustrative quotations.

**Table 4:** Barriers and Facilitators to GLA:D^®^ Implementation in Malawi mapped to the Consolidated Framework for Implementation Research.

| **INNOVATION DOMAIN** (GLA:D® program) |  |  |
| --- | --- | --- |
| **THEME 1:** The evidence-based content and organized structure of the GLA:D® program were perceived very positively by Malawian physiotherapists. Costs associated with implementing and day-to-day operation of the program were the main barriers identified. |  |  |
| **Construct** | **Sub-themes** | **Quotes** |
| Innovation Evidence-Base | Evidence supporting the GLA:D® program (F) | “What was most informative was also the evidence, some of the things that we didn't really know that they are working... we'll be able to be really confident when we are explaining to our patients on the management that we're giving them...it has worked. Of course, yes, in different populations, but at least we know that [GLA:D®] was tested some way and it has good evidence.” |
| Innovation Relative Advantage | Education in GLA:D® program will empower patients (F) | “I think the education is the key part...we were educating our patients, but then maybe we weren't really going deeper, but then after attending this program, we have learned how we can handle them from the start to the end.” |
|  | GLA:D® program will improve continuity of care (F) | “Also a proper follow-up will be there compared to the normal how we usually do our clinics for OA patients.”<br>“A patient can be seen with one therapist today and the next session they are seen by another therapist, and we don't have the guidelines set up on how we manage the patient, so you may find that the other therapist would not progress the patient from where the previous therapists stopped, so the continuation of the care is not as structured as it is in the GLA:D® program.” |
| Innovation Design | Uniformity and organization of GLA:D® program (F) | “We know that if the GLA:D® program is to be run, there will be uniformity in the treatment of patients with osteoarthritis.”<br>“We can gladly appreciate is the organisation of the program itself where one is equipped to learn where to know where to start, and then how you can progress to the next and so on” |
| Innovation Cost | Lack of funds to implement and operate GLA:D® program (B) | “If you are to get the [GLA:D®] program and implement it, it still comes back to the funding.” |
| **OUTER SETTING DOMAIN** (Community, health system, country). |  |  |
| **THEME 2:** Numerous sociocultural and systems level barriers identified included the entrenched current medical management of OA and traditionally limited awareness of physiotherapy services. Yet, a recent shifting within the medical community for greater acceptance and referral to physiotherapy earlier in the OA disease process was also highlighted by some. |  |  |
| **Construct** | **Sub-Themes** | **Quotes** |
| Local Attitudes | Malawi health system prioritizes management of infectious diseases over chronic conditions like OA (B) | “The problem with our healthcare system is that we are more concerned on infectious diseases and not these non-infectious or non-communicable diseases. So sometimes it's a little hard to actually implement some of these things like the acceptance, starting from the government level, the hospital level, even to the patients themselves is a little lower, the acceptance rates...There are a lot of campaigns on immunizations. Even though the rate is quite high, as we were saying that we see a lot of osteoarthritis patients, but not a lot of campaigns are being done about it, but there are a lot of campaigns on maybe malaria...HIV and a lot, so that could be one of the challenges that we could meet even when we are implementing such programs.” |
|  | Lack of awareness or knowledge about physiotherapy by other health professionals and general public, (B) yet recent shift towards increased and earlier referral to physiotherapy (F) | "Because here, the physiotherapy is an infant profession, it's just growing. A lot of people have little knowledge about physiotherapy...it's new to them."<br>"Even to the other health workers, they also have to know when to refer the patient. When is the right time to refer for the patient?"<br>"I think that is changing recently. Previously when people go to the orthopaedic clinic, they were treated with their analgesics only and then they would be sent back, but then in the recent years we've seen a change that usually when one goes to the orthopaedic clinic, when they are treated with the medications, they are then referred to physiotherapy as well, so the number of referrals has increased in such a way that we would say that as of now, they are more likely to be referred for physiotherapy other than just being managed by the orthopaedic clinicians." |
|  | Medications are the cornerstone of OA management and referrals to physiotherapy usually made as last resort after painkillers fail (B) | "Majority of the OA patients are managed medically, like prescription of painkillers, that's all."<br>"Like my friends say that the other treatment levels are not an option for most of our patients here, so their last resort is usually physiotherapy."<br>"By the time they'll come for physio, the damage has already been done. Maybe now the deformity that you can see – you can't really do much about it, but they are coming now to you because they are now immune or their body system has adapted to painkillers, so they extended that. When they take the painkillers, the pain is still there. So they don't really have any other options, but to come to you." |
| Local Conditions | No surgical options for OA management in Malawi health system (F) | "In Malawi, we only have medication and physio as the options for managing osteoarthritis. We don't have surgery as an option because we don't have it, currently, we're not doing it. So usually most of the patients, even though they are referred late, but they will still come back to physio" |
|  | Physiotherapists are not well integrated into health system, especially rural areas (B) | "So for a typical Malawian setup, physio is something that is just being adopted, I would say. It hasn't really synced in well with the system here. So most of the times when they allocate a physio to a particular district hospital or health centre, it's something where it might even take time for them to be given space to practice. And if they're given space to practice, sometimes they're given just days. So you're only allowed to use the space, say, Wednesdays or Fridays, something like that. That's with the space."<br>"Most areas that are far do not really have a hospital or physiotherapists close by. That's the other thing. So they have to travel long distance to come and reach to their nearest hospital." |
|  | Medical aid may cover costs of GLA:D® for patients (F) | "In Malawi, it means a medical aid which is more common here so for it sometimes they might add the sessions. So I feel like for the private sector, it might be doable as long as the patient hasn't been in physio already with so many sessions."<br>"Some [patients] might use their medical aid because they come in as referrals from the doctors so they can use their medical aid and it means the medical aid would be paying for them." |
| **INNER SETTING DOMAIN** (public and private hospitals and clinics) |  |  |

| **THEME 3:** Malawian physiotherapists agreed that implementation of GLA:D® would be initially easier in private hospitals and clinics due to better access to infrastructure, equipment, and funding compared to public settings. Involving rehabilitation technicians in settings with few or no physiotherapists was suggested. |  |  |
| --- | --- | --- |
| **Construct** | **Sub-Themes** | **Quotes** |
| Structural Characteristics: Work Infrastructure | Lack of coordination of physiotherapists to provide continuity of care (B) | "A patient can be seen with one therapist today and the next session they are seen by another therapist, and we don't have the guidelines set up on how we manage the patient, so you may find that the other therapist would not progress the patient from where the previous therapists stopped, so the continuation of the care is not as structured." |
|  | Rehabilitation technicians can assist physiotherapists (F) | "[Rehabilitation technicians] also very much involved in the management of these patients, so it's better that they also have the necessary knowledge and skills and training to be able to better assist these patients as well." |
|  | One-on-one physiotherapy appointments is norm (B) | "Mostly, it's one-on-one and then we follow up from there."<br>"I've never tried to offer a group therapy. It is something that you have to now sit back and say, "How am I going to implement this at my institution?" |
|  | Private hospital has better infrastructure to run GLA:D® program (F); where as public hospital has infrequent follow ups and booking are not made at specific time (B) | "It will work better in the private sector because of the resources that we have."<br>"It's quite different in our public settings where you have a lot of patients and you can't manage to see them even once every week. So you need maybe to see them maybe once a month. If one is lucky, maybe once in a fortnight. So maybe follow up to be a challenge."<br>"We usually have so many patients that come in, we don't usually book a patient that come at this particular hour. We just book patients come on this particular day. So you might have maybe 50 patients in the waiting line, waiting for five or six therapists to attend to them." |
| Relational Connections | Limited interprofessional collaboration (B) | "[Patients] go to an orthopaedic clinician, you do his part and you refer them to a physio. The physio will do his part. If he feels there is more needed, then you just refer them back. So it is that kind of collaboration. But coming together to discuss how to manage is very different, especially in this type of case, because it is mostly outpatients departments. So it is more you go back and the GP will also send that one back. You don't really meet in between to discuss how best." |
| Available Resources: funding, space, materials, and equipment | Lack of human resources: Physiotherapists (B) | "Where I work, I'm the only physio. Yeah, the rest are rehabilitation technicians. So then being the only physio, I will have to attend to this clients. It's like another role on top of whatever I already do." |
|  | Private hospitals may some patients who can pay for GLA:D® (F) | "If the patient has been seen in a private hospital, some of them might be able to pay for their sessions." |
|  | Public hospitals lack of funding for resources, space, and equipment (B) | "Even the resources we wanna use [in public hospitals], it all comes back to funding."<br>Usually the physio clinics are not designed like they were built as to be clinics. They're just spare rooms that were given for the physios to use. So the design itself is the one that makes the space to be smaller."<br>"It's just basically a room and a bed, and then the therapist has to improvise on the type of equipment that they use." |

| **INDIVIDUALS DOMAIN** (people with OA)<br>**THEME 4.** People with OA in Malawi were perceived to have numerous barriers across capabilities (low literacy, knowledge, mobility), opportunities (lack of transport, connectivity), and motivation (fearful of pain, no cost expectations). Some facilitators related to opportunity (having private insurance) and motivation (accepting of program and health professional advice) were suggested. |  |  |
| --- | --- | --- |
| Construct Name | Sub-Themes | Quotes |
| Capability | Low literacy (B) | "Maybe the literacy of the patient, as most of the patients who attend the public hospitals usually, may be not that literate." |
|  | Lack of awareness of physiotherapy (B) | "Physiotherapy is something that is not widely-known in our communities" |
|  | Older individuals with limited mobility and vision (B) | "The patients that we see, are usually elderly, so which brings in that limitation as well, some are already having difficulties with their vision, difficulties with technology, and the family illness." |
| Opportunity | Lack transport (B) | "Transport issues to bring/ ferry the people to the place, to the hospital, could be one of the challenges. Because for one to show up for their appointment, sometimes people miss the appointment because they do not have transport to ferry them to the hospital."<br>"Most areas that are far do not really have a hospital close by. That's the other thing. So [patients] have to travel long distance to come and reach to their nearest hospital." |
|  | Private health insurance or medical aid to pay for GLA:D® (F) | "If the patient has been seen in a private hospital, some of them might be able to pay for their sessions and some of them might use their medical aid because they come in as referrals from the doctors so they can use their medical aid and it means the medical aid would be paying for them." |
|  | Lack of internet connectivity or devices (B) | "And network could be a constraint in the areas, so as much as [patients] could be very interested [in telehealth], but now that becomes a challenge, because of the network."<br>"it's very difficult considering the use of technology... talk of internet connectivity and the charges that are happening with it, so it would be very difficult to follow up on a patient, very, often." |
| Motivation | High acceptability of GLA:D® (F) | "Also like my friends say that the other treatment levels are not an option for most of our patients here, so their last resort is usually physiotherapy. So I think they will actually receive it very well with the education that is incorporated, they might understand in general, I don't think there will be major issues in accepting the program among our patients here." |
|  | Fearful of pain and low compliance with exercises (B) | "So they'll depend on the medication rather than opting for physiotherapy because they also run away from the pain that we are talking about that after finishing the exercises, you may feel the pain, which is normal within 24 hours"<br>"So I feel pain when I do exercises, so I better opt for painkillers." |
|  | Receptive to health professional advice (F) | "But when it comes to medical advice, whether coming from a GP or a surgeon or a physio therapist, people listen to them."<br>"If it's a treatment, they might listen to any of them. There might be value since it's coming from someone from the hospital." |

|  | Expectations for incentives or that program is free (B) | "But if we go into the public sector, that's why we already say that it's a challenge because people in the public sector they hope that they will have the treatment for free." |
| --- | --- | --- |
| **IMPLEMENTATION PROCESS DOMAIN** (activities and strategies that could be used to implement GLA:D®)<br>**THEME 5:** Possible facilitators of implementation in Malawi included further adaptations to the GLA:D® physiotherapist training and how the program is delivered; initiatives to increase awareness and education of other health professionals and the general public about OA management and GLA:D®; and tailoring payment schemes and scheduling appointments for patients. |  |  |
| Construct Name | Sub-Themes | Quotes |
| Teaming | Partnering and collaborating with department heads, hospital leadership, specialty clinics (F) | "Management of different hospitals, like the hospital directors, so that if we let them know, so that they can provide support to the program."<br>"For this to work, we also need to collaborate with, especially the GPs that are the first contact usually most of the times when a patient has pain, they will go to them. So maybe as a team, if we would want to implement it, we also want to find ways on how to approach those that are our first contact and also spent time with them, maybe also tell them what the GLA:D® program is all about and how we can help each other." |
| Tailoring Strategies | Organize payment scheme options with patients (F) | "Most of the medical schemes that are used here in Malawi, they have a quota to how many sessions someone is allowed to attend the physio. So maybe it could be a part of their planning to check how many slots available for them and then how much are they willing to top up. So maybe to liaise with them, to be fair enough, since this is gonna take quite some time how much are they going to be willing to top up on their scheme. So maybe half the price would make sense for most of them, unless my friend needs to add more maybe." |
|  | Scheduling: one-on-one and group delivery options (F) | "Some of them, yes, may not do well with the group setup, but most of them, for what we have seen, do so much well with one-on-one"<br>"Maybe if we can organize – it all depends on organization. We can maybe group patients. Let's say, where we were working, we have a lot of patients with osteoarthritis, so maybe we can try to group them." |
| Engaging | Educate GPs, orthopedic teams, and nurses using presentations, posters, and flyers (F) | "Education sessions one-on-one [with GPs] is the best because our doctor would ask whatever questions they have and it means it's either through giving them a printout or having posters"<br>"In other communities, health centers, a GP might not be available, but nurses are available, so the nurses might be able to suggest to the patients what they can do. In case maybe they have not been referred, but the nurses might advise them where to go with their problems."<br>"Also maybe to do a proper education with the clinical and the orthopaedic team so that they are aware of this program running so that if they get to see most of them they should be able to refer them as many as they can so that many can participate and benefit." |
| Engage Innovation Deliverers | Physiotherapists will like strong evidence base and good organization of GLA:D training course (F) | "The introduction itself, the background and the definition, it was very informative, especially about the paradigm shift and everything else. There was a couple of new information, that was very good...How the whole program was introduced, it sort of made us more interested to really dive into the [GLA:D®] itself." |
|  |  | "The availability of the guidelines which at whatever point you get to see the client, you know exactly where to start from and then to tailor it according to the patient." |
|  | Embed physiotherapist training in health system (F) | "If we provide the program into the health system, maybe if the program is now owned by the health system, and the health system wants to advance the management of osteoarthritis by training more physios. So that is the way, maybe, people get trained." |
|  | Offer variety of training course formats and locations to make increase accessibly to physiotherapists across country (F) | "Those that manage to attend [GLA:D® training] are the ones based in Blantyre or up here. ... if it is a Zoom meeting, it means almost each and every district you have representative to connect."<br>"But in case where the in-person might not be very doable, then I think blending in the Zoom and a few meet-up points for people to practice and train via video would be doable." |
|  | Subsidized course fee and transportation for physiotherapists (F) | "If there was a provision of transport for other therapists, because others, they haven't come in because of where they are working now. Maybe if they were closer by or maybe they had the resources, they would have tried that because others, they stay very far from where the training has been held." |
|  | Certificates for training completion (F) | "Is there any way of giving maybe certification to those that are GLA:D® therapists or something that we know each other, |
|  | Incorporate African specific content into course (F) | "You could also incorporate, in terms of content from maybe Africa, since we are talking about Malawi, which is in Africa. So maybe in other African countries, if you could come up with some published articles, or if at all they are there, that directly interact with, talk about the African population, maybe that also would be helpful as well." |
| Engage Innovation Recipients | Public awareness and education campaigns about GLA:D program and physiotherapy (F) | "Out there in the communities, there's still little information on the presence of physiotherapists in general, not only for this condition, osteoarthritis. So we can take advantage of maybe informing the community about the presence of physiotherapy, while at the same time talk about osteoarthritis as well."<br>"So in our setting, each and every community has a village head. So once in a while there's a meeting that is called that each and every member of the community has to attend. So in those gatherings is where they now present different issues that are coming from the government that they want the people to hear. Or sometimes when there's a funeral ceremony is where also a chief or a village head gets to speak to the community of whatever is happening in the government on health or other issues that they can hear." |
|  | Incentives for patients (F) | "Even the clients themselves, for their motivation might be sort of a challenge, so incentives – I've seen some of the programs implemented and well if there is an incentive." |
| Adapting | Offer paper data collection (F) | "I think the registration would work – I mean the paper-based one would work and then maybe you can have the therapist later on transfer it into an online database, something like that, so that it's much easier." |
|  | Make education materials more visual and translation of materials (F) | "Instead of a projector, we could incorporate in some flip charts just to illustrate some of the things, but I think something that is way more doable is the printouts where you are using some of the visual graphics that you already part of the, on the websites where you have given the links, so I think it could be quite easy to print out some of those, the visual images and then the infographics and pass them around."<br>"I think that since most of the content is in English, it would require translation." |
|  | Physiotherapists adapt exercise to equipment and settings (F) | “Instead of theraband, we can use different weights. Like So like a sand bag, so you tie around the ankle, and you measure the level of difficulty, the level of difficulty depending on the weights that you have put in, and then you move on with that.”<br>“Instead of the physio ball, you could use a chair. Of course, it will not provide as much challenging balance as much, but at least it is an improvisation that could still work when you're doing the pelvic lift. So most of the exercises, we went through them and looked at some of the equipment that we could use to achieve the similar effect.” |
|  | Less frequent patient sessions: Train caregivers and/or follow up with device (F) | “Maybe changing the frequency of the time when the patients would be coming to the hospital that would be the case for the public hospitals, but then I'm not sure if that would not compromise the effectiveness of the program. But then as we've looked at it, most of the challenge for the patient is coming in with the cost of them attending the clinic frequently. So maybe if we could teach the patients the exercises so that they can be doing more of it at home other than coming to the clinic every time maybe it would help.”<br>“And for the private, since most of them could access the internet and the like, I think the telehealth now, if we could blend it in with the telehealth and a few of the physical [in-person] visits would work.”<br>“If we could involve the caretakers so that they are also trained, and maybe that way could reduce the number of the frequency to visit the physio.” |
|  | Use Rehabilitation Technicians to assist with GLA:D delivery (F) | “The core content is not that too technical for [rehabilitation technicians] to understand. It's more practical, and those guys are also not that bad in the hands-on practice. So if we incorporate them, that would be a plus for the program.” |
|  | Run GLA:D as 6-week camp in community (F) | “Community-based rehabilitation, where it's run as a camp, but it's something that will need a lot of funding, because for you to host, to keep a physio in such a camp for six weeks, or say maybe weekly, supporting the physio in a certain catchment area, and supporting them weekly with transportation and other things to meet the people in their communities, is something that I've seen other organizations doing.” |

**Innovation Domain:** Content and organization of the GLA:D^®^ program was thought to facilitate implementation for patients; however, program costs associated with implementation and operation were flagged as a critical barrier (Theme 1).

**Outer Setting Domain**: Numerous sociocultural and systems level barriers were identified such as the entrenched current medical model of management for OA that prioritizes painkillers and the overall greater focus of the health system on infectious diseases compared to chronic disease management. Yet, some participants suggested a recent shifting within the medical community for greater acceptance and earlier referral to physiotherapy for OA (Theme 2).

**Inner Setting Domain:** Key barriers identified included public hospital challenges (less infrastructure, care coordination, and equipment), with all participants agreeing that GLA:D^®^ implementation would be easier in private settings. Rehabilitation technicians were suggested to assist with GLA:D^®^ program in settings with few or no physiotherapists (Theme 3).

**Individual’s Domain:** Participants perceived that people with OA would have numerous barriers to accessing GLA:D^®^ that pertained to capabilities (e.g. low literacy), opportunities (e.g. lack of transport), and motivation (fearful of pain). Conversely, individual facilitators identified included having private insurance and being open and accepting of the GLA:D^®^ program and health professional advice.

**Implementation Process Domain:** Various facilitators of GLA:D^®^ implementation in Malawi included further adaptations to physiotherapist training (e.g. incorporating African specific content) and the GLA:D^®^ program (e.g. flexibility in number of sessions and delivery mode including one-on-one, telehealth); increasing awareness and education of other health professionals and the general public about OA and GLA:D^®^; and tailored patient focused strategies (e.g. make education material more visual) (theme 5).

## DISCUSSION

This mixed methods formative evaluation aimed to understand the future feasibility of implementing an OMP in a LMIC. Quantitative surveys found that after training, physiotherapists improved their knowledge and confidence about OA management, practice guidelines, and delivering care. Qualitative focus groups highlighted multi-level local barriers and facilitators to target before moving forward with implementation. Integrated data indicates high satisfaction with the GLA:D^®^ training course among Malawian physiotherapists and generated suggestions for adapting the physiotherapist training course and the GLA:D^®^ program to the Malawian context.

Malawian physiotherapists had the largest improvements in knowledge post-training for the questions on weight loss, joint symptoms, and the benefits of exercise for OA outcomes. These questions all relate to first line treatment of hip and knee OA, thus are critical for physiotherapists to know.^30^ The results also show that prior to the GLA:D^®^ training, all participants reported positive beliefs about the evidence for exercise therapy, patient education, and weight management in OA management.

These positive beliefs may be related to the qualitative finding that there has been a recent shift within the Malawian medical community for earlier referral to physiotherapy and exercise therapy for OA.

Our participants reported the high use of medications (painkillers) and the predominantly biomedical approach for the management of OA in Malawi. Other African studies mirror this management approach, as pain medication has been found to be the mainstay of OA management in rural South Africa,^31^ while in Nigeria only 11% of physiotherapists used therapeutic exercise as a treatment modality.^32^ This may be because physiotherapists are often not the first point of contact for patients in Africa, with physiotherapist-led OA care often being difficult to access, and considered unaffordable (needing to be paid for out of pocket).^6^ Hence, while our participants acknowledged the importance of these domains in the first line management, they seldom apply them in practice. Globally, despite ample evidence showing the efficacy of first line treatments in improving outcomes in OA, the gap between best evidence and practice persists.^8,30,33^

After training, Malawian physiotherapists reported improved confidence in delivering exercise therapy and education to people with hip and knee OA and improved confidence to follow clinical practice guidelines. Ideally, this will translate to improved patient care, as has been previously found in studies involving general practitioners and nurses.^34,35^ These studies found improved knowledge after engagement with learning activities such as internal in service training or courses led to clinicians improving the quality of care for their patients and increasing uptake of clinical practice guidelines. However, implementing new knowledge and improving clinical outcomes for patients with OA in Malawi, as in other LMICs, may be challenging as rehabilitation professionals and practice are not yet well integrated or prioritised into the health system.^36^ This was echoed in our qualitative findings, where participants highlighted the difficulty in implementing OA rehabilitation programs with minimal governmental support. Equally neglected is the training of rehabilitation professionals, development and implementation of treatment guidelines, and coordination of care.^6,37,38^ Therefore, as suggested by participants in the focus groups, successful implementation of OMP like GLA:D^®^ in LMICs will not be facilitated by training physiotherapists and other rehabilitation professionals (e.g., rehabilitation technicians) alone. Raising awareness among other healthcare professionals, including nurses and physicians, and the general public on the effectiveness of exercise-based interventions in the management of OA is also needed.

Malawian physiotherapists’ confidence in delivering education about total joint replacement and arthroscopic surgery was largely unchanged post-course. This is unsurprising as our participants qualitatively reported that joint replacement surgery is not available in Malawi. Hence, physiotherapists do not routinely discuss it with their patients. The global demand for joint replacement surgery is on the rise and predicted to further increase in high income countries.^39,40^ Consequently, desiring or receiving surgery is commonly reported in studies on OA in high income countries, and physiotherapists would have discussions about these procedures with their patients.^15,19,41^ In contrast, joint replacement procedures in LMICs are rare as limited orthopaedic services in these health systems prioritise traumatic injuries or orthopaedic complications of HIV infection and/or antiretroviral treatment.^42–44^ Going forward, the OMP content related to joint replacement surgery should be contextually adapted to be relevant to LMICs.

Our findings are similar to previous research examining GLA:D^®^ training outcomes in physiotherapists. Malawian and Australian physiotherapists both showed improved confidence in knowing how to deliver exercise and education, as well as confidence in their ability to deliver exercise and education following clinical practice guidelines.^45^ Also reflective of our findings, the structure and organisation of the GLA:D^®^ program, buy in from other health care providers, and a supportive organisational context were identified as key enablers to the implementation of the program by physiotherapists in Switzerland, Australia, and Canada.^45–47^ Malawian physiotherapists perceived the challenges associated with GLA:D^®^ implementation to be greater in public hospitals, with implementation likely to be easier in private settings. The same was observed in Australia, where only 15% of physiotherapists and 11% of implementation sites in the GLA:D^®^ Australia program are from public settings.^19^ Personal cost to patients, especially those with low incomes, was also identified as a potential barrier to implementation by rehabilitation professionals in Australia and Canada, similar to our sample.^46^ Our qualitative findings further suggested that rural settings may pose additional challenges in the Malawian context, such as lack of transportation and no scheduling of appointments. Similarly, research in Canada and Australia found that the GLA:D^®^ program was not easily accessible in rural settings. However, when accessed, the program led to comparable outcomes in patients from urban or rural settings.^19,48^

In our focus groups, participants suggested that telehealth may be a viable option for facilitating physiotherapist follow up, while reducing transportation burden. GLA:D^®^ delivered by telehealth is reported to be acceptable and provides similar outcomes as in-person OA management.^49–51^ However, barriers to telehealth adoption such as computer literacy, internet connectivity, and equipment access have been noted even in high income countries.^49,50^ Additionally, prior experience with telehealth influences its acceptability among OA patients.^49^ Notably, South African patients with OA were sceptical of telerehabilitation programs as they lack physical interaction, although they acknowledge the positives such as accessibility, and convenience.^52^

The study findings should be considered in the context of some limitations. Firstly, this formative evaluation engaged with 11 Malawian physiotherapists in one region (Blantyre), which may limit the transferability of our results to other Malawian physiotherapists and other LMICs. However, this geographical restriction was necessary to ensure physiotherapists could come to Kamuzu University of Health Sciences for the GLA:D^®^ training course to mitigate potential internet connectivity issues. This evaluation focused on physiotherapists’ perspectives of facilitators and barriers to implementation of the GLA:D^®^ program. Inclusion of other health professionals and patients with OA in future studies may provide additional insights. Future work should not only evaluate clinical outcomes of OMP such as GLA:D^®^ but also consider expanding to examine the role of other health professionals and patients.

## CONCLUSION

Our study found that the GLA:D^®^ training course improved Malawian physiotherapists’ overall knowledge of first line OA treatment and confidence in delivering exercise-therapy and patient education. High use of pain medications and the predominantly biomedical approach for the management of OA in Malawi was also reported reflecting the fact that physiotherapists are rarely the first point of contact for patients in the Malawian context. Costs associated with program set up and delivery, and lack of transportation for patients were reported as key implementation barriers, especially in public health settings. Increasing education and awareness of physiotherapists, other health professionals, and the general public about evidence-based OMP such as GLA:D^®^, as well as making contextual adaptions to the these programs (e.g., adding African specific content, offering transport) and program structure (e.g., less in-person sessions, use of rehabilitation technicians) may facilitate future implementation of OMP in LMICs.

## KEY POINTS

- Findings

- After completing the GLA:D^®^ training, Malawian physiotherapists had improved knowledge and confidence in delivering first line treatment (exercise-therapy and education) for hip and knee OA. They were highly satisfied with the GLA:D^®^ training, and suggested numerous strategies for adapting the training and the program to a LMIC context.
- Physiotherapists revealed key multi-level barriers for implementation including culturally entrenched medical management of OA, lack of clinic funding and resources, and low literacy and no transport for patients. However, they also highlighted potential facilitators including increased acceptance of exercise by public, use of rehabilitation technicians, and increasing awareness, education, and collaborations with other health professionals.

- Implications

- This formative evaluation generated valuable knowledge that can inform future implementation of OMP in LMICs. The GLA:D^®^ training and the GLA:D^®^ program should be adapted to account for key systems and person level barriers and leverage the facilitators that are unique to LMICs.

- Caution

- This preliminary study included a small number of physiotherapists practising in one urban region of Malawi. The results should be interpreted with caution and viewed as the first step to implementing a OMP in a LMIC.

## SUPPLEMENTARY TABLES

Supplementary Table 1: Focus group structure and questions

Supplementary Table 2: Pre and post-GLA:D^®^ training course OA knowledge and management among Malawian physiotherapists (n=11)

Supplementary Table 3: Malawian physiotherapists post-GLA:D^®^ training knowledge of GLA:D^®^ procedures and practices (n=11)

## Data Availability

All data produced in the present study are available upon reasonable request to the authors

## Acknowledgements

We would like to acknowledge the Physiotherapy Association of Malawi for their assistance in participant recruitment, the physiotherapists who participated in the study, as well as GLA:D^®^ Australia leadership for their in-kinds support to the study.

## Author contributions

NSM – Study conceptualisation, Data analysis, Manuscript writing

EJ – Participant recruitment, Data collection, Manuscript writing

EMC – Study conceptualisation, Participant recruitment, Data collection, Manuscript writing

CB – Study conceptualisation, Data collection, Manuscript writing

JLK – Study conceptualisation, Data collection, Manuscript writing

AE– Study conceptualisation, Data collection, Data analysis, Manuscript writing

All the authors provided critical appraisal of the manuscript and approved the final version of the manuscript before submission.

## Ethics approval

The study was approved by the Kamuzu University of Health Sciences, College of Medicine Research and Ethics Committee – Malawi (REF: P.08/22/3707) and La Trobe University Human Research Ethics Committee – Australia (REF: HEC23003)

## Data sharing statement

Individual, de-identified data collected in the study will be made available following publication, for aims approved in the proposal and upon reasonable request to the corresponding author on. These data will be available for 3 years after publication.

