## Supplementary tables for "Supporting the implementation of Osteoarthritis Management Programs in low-and middle-income settings: Understanding clinician training outcomes alongside perceived program barriers and facilitators in Malawi"

**APPENDIX D**

**GLA:D adaptation for Malawi PT workshop agenda**

**(Date TBD)**

| Time | Item | Leads (TBC) |
|---|---|---|
| 10 mins | Welcome to event, introduce facilitators<br>Re: confirm recording<br>Set expectations:<br>that all information is confidential and dealt with in a respectful manner. Should participants feel uncomfortable at any point during the workshop they are free to private message the facilitator and withdraw. | AE |
| 5 mins | Introduction to tasks | AE |
| 20 mins | Break out room activity 1 (Intervention characteristics & characteristics of individuals)<br><br>*Potential adaptations to GLA:D training course and GLA:D program*<br>*Understanding physiotherapists and patients with OA in Malawi* | 3 facilitators |
| 15 mins | Large group discussion from breakout rooms 1 | AE |
| 15 mins | Breakout room activity 2 (Inner & outer settings)<br><br>*Patient needs/resources, role of other health professionals, culture*<br>*Understanding different settings (hospital vs clinic)* | 3 facilitators |
| 15 mins | Large group discussion from breakout rooms 2 | AE |
| 10 min | BREAK | |
| 15 min | Break out room activity 3 (Process)<br><br>*Planning to implement GLA:D*<br>*Engaging Stakeholder groups* | 3 facilitators |
| 15 mins | Large group discussion from breakout rooms 3 | AE |
| 5 mins | Close/summary/next stages | AE |

**BREAKOUT 1 (20 min): Intervention characteristics & characteristics of individuals**

- How do you think the GLA:D training course should be adapted to meet the needs for physiotherapists in Malawi?

  Prompt: How interested do you think physiotherapists will be to take the GLA:D program? Potential barriers/enablers to physiotherapists taking the GLA:D training?

- How do you think the GLA:D program should be adapted to be successfully provided to patients with osteoarthritis in Malawi?

Approved by
College of Medicine
19-Oct-2022
(COMREC)
Research and Ethics Committee

- Please describe how people with osteoarthritis are usually managed your region?

- How do you think patients will perceive the GLA:D program? (i.e. value, cost, complexity)

<u>**BREAKOUT 2 (15 min):**</u> **Inner & outer settings**

- What contextual factor (e.g. system, political, cultural) should be considered when developing strategies to implement GLA:D in Malawi?

- Where would you envision patient most likely to access GLA:D (i.e. hospital, private clinic) and why?

Prompt: What role would you see other health professionals having with GLA:D? (i.e. GP referral, ortho wait lists).

- How would you describe the perceived need for a program like GLA:D at your hospital or clinic?

Prompts: How would you see it fitting in with current care provided? What would you see as potential barriers? Can you describe any possible enabling factors?

<u>**BREAKOUT 3:**</u> **Process**

- What challenges and opportunities are there for implementing GLA:D in Malawi?
- What do you think would be the most important step(s) to prepare to implement the GLA:D program at your hospital/clinic?

- Who do you think we need to engage with to help with successful implementation? (i.e. health care administrators, community leaders, etc)

How to you see these stakeholders assisting with implementing GLA:D?

Approved by
College of Medicine
19-Oct-2022

(COMREC)
Research and Ethics Committee

**Supplementary Table 2: Pre- and post-GLA:D® training course OA knowledge and management among Malawian physiotherapists (n=11)**

| **Questions/Responses** | **Pre-GLA:D® training frequency**<br><br>**n (%) correct** | **Post-GLA:D® training frequency**<br><br>**n (%) correct** | **Effect size**<br><br>**(p-value)** |
|---|---|---|---|
| Which of the following statements is also true for osteoarthritis?<br>Osteoarthritis is a joint disease which in addition to joint pain may cause psoriatic arthritis<br>**Osteoarthritis is a joint disease which may affect a single joint or multiple joints**<br>Osteoarthritis is a symmetric joint disease requiring medical treatment<br>Osteoarthritis is a joint disease which manifests as inflammation of the big toe | 11 (100) | 11 (100) | – |
| **Which three elements are placed at the bottom of the pyramid indicating first-line treatment for knee/hip osteoarthritis?**<br>Exercise, patient education and medication<br>**Exercise, patient education and weight loss (where relevant)**<br>Exercise, orthoses, and weight loss (where relevant)<br>Exercise, manual treatment and patient education | 6 (54.5) | 11 (100) | 0.45 (0.063) |
| **Based on which factor(s) are you able to make the clinical diagnosis of osteoarthritis?**<br>**The diagnosis can be made based on an overall assessment of symptoms, clinical findings and risk**<br>The diagnosis can be made based on clinical findings and risk factors<br>The diagnosis can be made based on the symptoms alone<br>The diagnosis can only be made based on an x-ray | 11 (100) | 11 (100) | – |
| **Which of the following diagnoses is least likely to be seen among people with osteoarthritis?**<br>**Rheumatoid arthritis**<br>Depression<br>Diabetes<br>Hypertension | 6 (54.5) | 7 (63.6) | 0.09 (1.000) |
| **Which joints are most often affected by osteoarthritis?**<br>Hip knee and ankle joints<br>**Hip knee and finger joints**<br>Hip knee and elbow joints | 4 (36.4) | 8 (72.7) | 0.36 (0.125) |

| | | | |
|---|---|---|---|
| Hip knee and shoulder joints | | | |
| **Does the intensity of pain at baseline or the severity of radiological findings impact on whether someone with osteoarthritis can benefit from therapeutic and the exercise?**<br>Yes, the degree of radiographic osteoarthritis has an impact on the effect of exercise<br>Yes, both pain intensity at baseline and the degree of radiographic osteoarthritis have an impact on the effect of exercise<br>Yes, pain intensity at baseline has an impact on the effect of exercise<br>**No, people with osteoarthritis can benefit from therapeutic exercise regardless of pain or radiographic severity** | 1 (9.1) | 11 (100) | 0.91 (0.002) |
| **What do the clinical guidelines on knee osteoarthritis say about the nutritional supplement glucosamine?**<br>**It may have a small positive effect for some people**<br>It is dangerous for people with diabetes<br>The research has come from reliable sources<br>People with osteoarthritis should take glucosamine, even if they don't feel it is helping them | 7 (63.6) | 11 (100) | 0.36 (0.125) |
| **Which of the following statements does NOT fit the description of articular cartilage?**<br>**The volume of articular cartilage may be rebuilt through correct exercise**<br>Synovial fluid diffuses into and out of the articular cartilage carrying with it a supply of fit the nutrients<br>Articular cartilage reduces friction during movement<br>There are no nerves in cartilage tissue, so it does not produce pain | 6 (54.5) | 6 (54.5) | 0.00 (1.000) |
| **What is the duration of morning stiffness in patients with osteoarthritis according to clinical guidelines for osteoarthritis?**<br>10 minutes<br>**30 minutes**<br>5 minutes<br>120 minutes | 4 (36.4) | 6 (54.5) | 0.19 (0.687) |
| **What do the RACGP guidelines recommend about long-term use of oral NSAIDs for someone with cardiovascular disease?**<br>**They advise against it**<br>They recommend it for patients who have difficulty walking<br>They recommend it for patients suffering from swelling<br>They recommend it for patients experiencing nightly pain | 6 (54.5) | 5 (45.5) | 0.1 (1.000) |

| | | | |
|---|---|---|---|
| **Certain objective findings in knee osteoarthritis patients clearly indicate osteoarthritis - which are they?**<br>Positive drawer test, hypermobility, and joint line tenderness<br>Muscle soreness in calf, inner thigh, and thigh<br>Swelling, extension defect and quadriceps tender to palpation<br>**Crepitation, reduced range of movement and possibly bone thickening on palpation** | 10 (90.9) | 11 (100) | 0.09 (1.000) |
| **How much of a weight loss is recommended for overweight knee osteoarthritis patients to achieve pain reduction and improved function?**<br>At least 15%<br>**At least 5%**<br>At least 2%<br>At least 20% | 1 (9.1) | 10 (90.9) | 0.82 (0.004) |
| **Which of the following statements are NOT considered a risk factor for the development of osteoarthritis?**<br>**Physical exercise**<br>Overweight<br>Previous joint injury<br>Quadriceps muscle weakness | 9 (81.8) | 10 (90.9) | 0.1 (1.000) |
| **How does worsening or flare-up usually behave if the GLA:D exercise program is continued?**<br>It will invariably fluctuate<br>**It will gradually reduce**<br>It will gradually increase<br>It remains unchanged over time | 9 (81.8) | 6 (54.5) | 0.27 (0.453) |
| **For how many hours is it acceptable to feel a worsening of symptoms after a training session?**<br>72 hours<br>**24 hours**<br>12 hours<br>48 hours | 2 (18.2) | 10 (90.9) | 0.73 (0.008) |
| **Which risk factors for osteoarthritis are NON-modifiable?**<br>Age, height, and hereditary predisposition (genetics)<br>Age, sex, and weight<br>Age, height, and weight<br>**Age, sex, and hereditary predisposition (genetics)** | 9 (81.8) | 11 (100) | 0.18 (0.500) |
| **What is generally characterized as acceptable pain on the VAS scale during a training session?** | 8 (72.7) | 11 (100) | 0.27 (0.250) |

| | | | |
|---|---|---|---|
| 2<br><br>**5**<br><br>10<br><br>7 | | | |
| **According to the RACGP guidelines, which type of painkiller in the following list is the safest choice for pain relief for osteoarthritis?**<br><br>Oral NSAIDs/ Topical NSAIDs/ Paracetamol | 11 (100) | 11 (100) | – |
| **Which three features are key signs of osteoarthritis?**<br><br>- Increased pain and stiffness when you have not moved your joints for a while, worsening with activity, easing with stretching<br>- **Increased pain and stiffness when you have not moved your joints for a while, easing with movement and worsening with increased load**<br>- Increased pain and stiffness when you have not moved your joints for a while, worsening with non-weight-bearing movement and easing with weight-bearing activity<br>- Increased pain and stiffness when you have not moved your joints for a while, worsening with activity and easing with prolonged walking | 10 (90.9) | 9 (81.8) | 0.09 (1.000) |
| **Which of the following persons are NOT likely to have osteoarthritis?**<br><br>- A 25-year old woman who has had an ACL reconstruction in her teens<br>- An overweight middle-aged woman<br>- **A 17-year old girl with persistent and inexplicable pain**<br>- An elderly person with swelling and rest pain | 9 (81.8) | 9 (81.8) | 0.00 (1.000) |
| **Which of the following persons are NOT likely to have osteoarthritis?**<br><br>- A 58-year old woman with long-lasting load-dependent pain<br>- **A 45-year old man with pain along the outside of the knee after a period of progressively intensifying running**<br>- A 41-year old overweight woman who is experiencing pain while walking<br>- A 68-year old man suffering from brief morning stiffness and who has problems climbing stairs | 10 (90.9) | 10 (90.9) | 0.00 (1.000) |

### Supplementary Table 3: Malawian physiotherapists post-GLA:D® training knowledge of GLA:D® procedures and practices (n=11)

| Question/Responses | n (%)<br>correct response |
|---|---|
| **What are the overall objectives of neuromuscular exercises?** | |
| **Improved sensory-motor control and functional joint stability** | 11 (100) |
| Improved movement and balance | |
| Improved static strength and fitness | |
| Improved endurance and eccentric strength | |
| **Based on resistance training principles, neuromuscular exercises are most likely to improve:** | |
| Muscular atrophy | |
| **Muscular strength endurance** | 10 (91) |
| Muscular length | |
| Muscular power | |
| **In the two sliding exercises that help with controlling dynamic alignment, which leg is the weight bearing leg in this exercise?** | 11 (100) |
| **The leg without the sliding material under the foot** | |
| The leg with the sliding material under the foot | |
| The patient must stand with equal weight bearing on both feet | |
| The weight shifts from leg to leg | |
| **Which of the following statements has NO influence on the physiotherapist's decision to progress the exercise in the GLA:D program?** | |
| The physiotherapist's visual assessment of quality of the movement | 10 (90.9) |
| No increase in pain in the 24 hours following a exercises training session | |
| Pain rated below 5 on the VAS scale during exercise | |
| **The findings from a new x ray report provided by the patient** | |
| **What is the proper knee position during the chair stand exercise?** | |
| **The same distance between the knees throughout the entire exercise** | 10 (91)** |
| The knees should move inwards when the patient sits down | |
| The knees should move outwards both on the way up and the way down | |
| The knees should move inwards when the patient gets up | |
| **How should the GLA:D exercise program be adjusted to patients with only one osteoarthritis-affected joint?** | 9 (82) |
| The affected leg is trained twice as hard as the basis and adapted to the functional level of each non-affected leg patient | |

| | |
|---|---|
| Only the affected leg is trained<br>**Both extremities are trained approximately equally**<br>Each exercise involving the affected leg includes five additional repetitions | |
| **Which of the following statements is most INCORRECT when considering the dynamic alignment of hip, knee, and foot during a one stance exercise such as the step up or lunge exercises?**<br>**Alignment is not important**<br>The patient should focus on proper alignment of the non-weight-bearing leg<br>The patient should focus on proper alignment of the weight-bearing leg<br>The patient should focus on good alignment of both the stance leg and the exercise leg (weight-bearing and non-weight-bearing leg) | 8 (73) |
| **What is the prescription for the number of repetitions and sets that form the basis of each exercise in the GLA:D program?**<br>4-5 sets of 10-15 repetitions<br>4-5 sets of 5-8 repetitions<br>**2-3 sets of 10-15 repetitions**<br>2-3 sets of 5-8 repetitions | 10 (91) |
| **What is involved in the 12-month post GLA:D program follow-up?**<br>Patient questionnaire and physical tests<br>Patient questionnaire and physiotherapist form<br>Consultation with physiotherapist with a view to continued exercise participation<br>**Only patient questionnaire automatically sent to the patient's e-mail address** | 7 (64) |
| **Which physical tests are mandatory and which physical test is optional in GLA:D®?**<br>Timed-up-and-go (TUG) and 40m walk test are mandatory and 30 seconds chair stand test is optional<br>A Timed-up-and-go (TUG) and 6 minutes walk test are mandatory and 40m walk test is optional<br>**40m walk test and 30 seconds chair stand test are mandatory and single-leg hop test is optional**<br>6 minute walk test and 30 seconds chair stand test are mandatory and single-leg hop test is optional | 9 (82) |
| **Which of the following patients should you include in GLA:D®?**<br>**A patient who is already physically active**<br>A patient who is unable to understand and communicate in English<br>A patient with other symptoms that are more pronounced than the osteoarthritis problems<br>A patient whose symptoms are due to other problems than osteoarthritis | 9 (82) |
| **In order to provide the GLA:D® program what 3 elements must you offer?**<br>Patient education, 6 weeks of neuromuscular exercise and physical activity training | 8 (73) |

| | |
|---|---|
| Patient education, 6 weeks of neuromuscular training sessions and weight loss advice<br><br>**Patient education, 6 weeks of neuromuscular training sessions and outcome measures**<br><br>Patient education, weight loss and physical activity training instructions | |
| **What is the main focus of neuromuscular exercises?**<br><br>That the patient gets his or her heart rate up<br><br>That the patient is able to perform each exercise as quickly as possible<br><br>**That the patient performs the exercises with focus on controlling their alignment**<br><br>That the patient is able to do as many repetitions as possible | 8 (73) |
| **How should you adjust the neuromuscular exercise program to the individual patient?**<br><br>**Use the different levels of difficulty built into the program**<br><br>Those with a high functional level should do the quality exercise program three times a week<br><br>Those with a high functional level and those with a low functional level should do entirely different exercise programs<br><br>Those with a low functional level should only do half of the exercises in the program | 10 (91) |
| **When should the post GLA:D program follow-up take place?**<br><br>Three months after the last GLA:D® training sessions<br><br>**Three months after initial consultation and registration**<br><br>Three months after the first GP consultation<br><br>Three months after the symptoms began | 6 (55) |

** One (1) missing response; Correct answers are in **bold**

Supplementary Table 5: Completed **consolidated criteria for reporting qualitative studies (COREQ): 32-item checklist**

| **No. Item** | **Guide questions/description** | **Reported on Page #** |
|---|---|---|
| **Domain 1: Research team and reflexivity** | | |
| *Personal Characteristics* | | |
| 1. Inter viewer/facilitator | Which author/s conducted the interview or focus group? | 10 |
| 2. Credentials | What were the researcher's credentials? E.g. PhD, MD | 10 |
| 3. Occupation | What was their occupation at the time of the study? | 10 |
| 4. Gender | Was the researcher male or female? | 10 |
| 5. Experience and training | What experience or training did the researcher have? | 10 |
| *Relationship with participants* | | |
| 6. Relationship established | Was a relationship established prior to study commencement? | **11** |
| 7. Participant knowledge of the interviewer | What did the participants know about the researcher? e.g. personal goals, reasons for doing the research | - |
| 8. Interviewer characteristics | What characteristics were reported about the inter viewer/facilitator? e.g. Bias, assumptions, reasons and interests in the research topic | - |

| | | |
|---|---|---|
| **Domain 2: study design** | | |
| *Theoretical framework* | | |
| 9. Methodological orientation and Theory | What methodological orientation was stated to underpin the study? e.g. grounded theory, discourse analysis, ethnography, phenomenology, content analysis | 10 |
| *Participant selection* | | |
| 10. Sampling | How were participants selected? e.g. purposive, convenience, consecutive, snowball | 8 |
| 11. Method of approach | How were participants approached? e.g. face-to-face, telephone, mail, email | 8 |
| 12. Sample size | How many participants were in the study? | 8 |
| 13. Non-participation | How many people refused to participate or dropped out? Reasons? | - |
| *Setting* | | |
| 14. Setting of data collection | Where was the data collected? e.g. home, clinic, workplace | 8 |
| 15. Presence of non-participants | Was anyone else present besides the participants and researchers? | - |
| 16. Description of sample | What are the important characteristics of the sample? e.g. demographic data, date | 13 |
| *Data collection* | | |
| 17. Interview guide | Were questions, prompts, guides provided by the authors? Was it pilot tested? | 10 |
| 18. Repeat interviews | Were repeat inter views carried out? If yes, how many? | **n/a** |
| 19. Audio/visual recording | Did the research use audio or visual recording to collect the data? | 11 |
| 20. Field notes | Were field notes made during and/or after the inter view or focus group? | no |
| 21. Duration | What was the duration of the inter views or focus group? | 10 |
| 22. Data saturation | Was data saturation discussed? | no |
| 23. Transcripts returned | Were transcripts returned to participants for comment and/or correction? | no |
| **Domain 3: analysis and findings** | | |
| *Data analysis* | | |
| 24. Number of data coders | How many data coders coded the data? | 11 |

| | | |
|---|---|---|
| 25. Description of the coding tree | Did authors provide a description of the coding tree? | No |
| 26. Derivation of themes | Were themes identified in advance or derived from the data? | 11 |
| 27. Software | What software, if applicable, was used to manage the data? | **11** |
| 28. Participant checking | Did participants provide feedback on the findings? | no |
| *Reporting* | | |
| 29. Quotations presented | Were participant quotations presented to illustrate the themes/findings? Was each quotation identified? e.g. participant number | Supplementary Table 4 |
| 30. Data and findings consistent | Was there consistency between the data presented and the findings? | Yes |
| 31. Clarity of major themes | Were major themes clearly presented in the findings? | Yes |
| 32. Clarity of minor themes | Is there a description of diverse cases or discussion of minor themes? | Table 4 |
